# The intention to use Kangaroo Mother Care among first-time mothers 15-24 years in Kinshasa, DRC: An application of the Integrated Behavioral Model

**DOI:** 10.64898/2026.02.04.26345616

**Authors:** Madeline Woo, Anastasia J. Gage, Julie Hernandez, Pierre Z. Akilimali, Jane T. Bertrand

## Abstract

**Background:** Kangaroo mother care (KMC) is a low-cost, safe, and simple method that has proven efficacious at improving neonatal health outcomes, including mortality, in low-income countries. While beneficial for all newborns, the World Health Organization (WHO) recommends KMC specifically for low birthweight (LBW) and premature infants. This study, guided by a modified Integrated Behavioral Model (IBM) framework, examined whether attitudes, norms, and personal agency predicted KMC intentions among first-time mothers (FTMs) 15-24 years old in Kinshasa, the Democratic Republic of Congo (DRC).

**Methods:** This study used secondary data from the 2020 Momentum Project endline survey, a quasi-experimental design in three intervention and three comparison health zones in Kinshasa, DRC. The association between IBM components and KMC intentions was tested using multivariate logistic regression. The total sample consisted of 1,918 adolescent and young first-time mothers (FTMs) from six health zones.

**Results:** About half of the respondents stated that they were very likely to practice KMC if they had a LBW or preterm baby. Approval of KMC, normative expectations, descriptive norms, self-efficacy, and autonomy were all positively associated with the intent to practice KMC. Age was also found to be a significant predictor; the odds of KMC intentions were 90% higher (95% CI=(1.11-3.24)) for women 20-24 years compared to women 15-19 years. The association between autonomy and KMC intentions was moderated by age. Marginal effects of the interaction found that the probability of intending to practice KMC for respondents 20-24 years was constant regardless of autonomy level, while autonomy significantly increased the probability of KMC intentions for younger women 15-19 years.

**Conclusions:** Results suggest that KMC intentions among first-time mothers 15-24 years are influenced by multiple factors simultaneously. Programs seeking to improve KMC intentions among this population should consider implementing age-specific empowering and norms-based strategies.

## Introduction

Neonates (0-28 days old) make up the largest share of under-five deaths worldwide (44%) [1], with three-fourth of deaths occurring during the early neonatal period (days 0-6) [2]. Complications from preterm birth is the leading cause of neonatal death globally [3] and Sub-Saharan Africa has the highest rate of neonatal mortality [4,5]. Located in Central Africa, the Democratic Republic of Congo (DRC) has the 24^th^ highest neonatal mortality rate [5] in the world (27 deaths per 1,000 live births [6]) and is one of five countries that contribute to nearly 50% of all neonatal deaths worldwide [7]. The DRC has a preterm birth rate of 10 births <37 weeks per 100 live births [8] with a low birthweight (LBW) (<2500 grams) prevalence of 11% [9], and preterm birth complications are the leading cause of neonatal death (42%) [10].

First endorsed by the World Health Organization (WHO) in 2003[11], Kangaroo mother care (KMC) is a method developed to promote well-being among LBW and preterm neonates [12]. KMC is comprised of prolonged skin-to-skin contact between the newborn and caregiver, started as quickly as possible after birth and practiced continuously for as long as possible (ideally 24 hours), exclusive breastfeeding, and early discharge with follow-up care [12–14]. The clinical effectiveness of KMC in reducing neonatal mortality and morbidity, including in the first week of life [15,16], has been demonstrated in numerous settings, including low-resource settings [14,17].

One population that could especially benefit from KMC are the newborns of adolescent (15-19 years) and young (20-24 years old) mothers. The WHO estimates that between 14 to 15 million adolescent girls give birth each year; about 10% of all births worldwide, with over 90% of these births taking place in low- and middle- income countries (LMICs) [18]. Young maternal age is associated with adverse health outcomes in infants including LBW, preterm birth, and neonatal mortality [19–22]. Newborns of adolescents have a 50% risk of stillbirth or dying before one month in comparison to newborns of mothers 20-29 years [23]. Globally, Sub-Saharan Africa has the highest fertility rates for adolescent (101.0 births per 1,000 women) and young women (202.9 births per 1,000 women) [24]. The fertility rate for adolescent and young women in the DRC is higher than the regional average at 109 births per 1,000 women 15-19 years and 250 births per 1,000 women 20-24 years [25] respectively. Research focused on KMC among adolescent and young mothers is limited, especially in low and middle-income countries (LMICs), with only one qualitative study conducted in South Africa examining young mothers’ lived experiences with KMC [26].

### KMC in the DRC

KMC initiatives have not been widely implemented in the DRC and studies have focused primarily on policy, facility implementation, and provider knowledge, not KMC-related attitudes, perceptions and norms. In 2012, the government adopted a policy on standards of maternal, child, adolescent and newborn health which advocated for prolonged skin-to-skin contact for premature and LBW neonates [27]. In 2018, the DRC updated its policy/guidelines on KMC to include an indicator for newborns who benefited from KMC in the health management information system (HMIS) [28].

Based on available documentation, KMC has been implemented in the DRC primarily through two settings: the United States Agency for International Development’s (USAID) multi-sectoral Integrated Health Project (IHP) and a tertiary hospital in Lubumbashi, DRC. From 2010-2015 the IHP conducted trainings on KMC at health facilities in four southwest provinces as one of several interventions to improve maternal, child, and newborn health (MCNH) indicators in the DRC [29,30]. A quasi-experimental retrospective study at an IHP implementation hospital in Buta, DRC found a statistically significant decrease in the odds of neonatal survival in the 36 months after program implementation compared to the 36 months prior [31].

According to personal communications with Dr. Narcisse Embeke, the deputy chief of party for IHP in the DRC, a new phase of IHP began in 2018 in nine of the same ten provinces in which the program was previously implemented, and from 2018-2020 IHP provided supervision of health facilities that had already integrated KMC. Starting in 2021, IHP began providing training and refresher training to providers on KMC, extended KMC trainings to new health facilities, and gave health facilities materials and equipment to support KMC such as infant wraps. The program did not, however, work directly with the community to increase awareness of or interest in KMC.

A separate KMC program was established in 2013 at a tertiary hospital in Lubumbashi, DRC. A cohort study of the program found that only 39% of qualifying neonates received KMC [32]. The low rate of utilization was due to a lack of knowledge about KMC among both healthcare workers and caregivers; however, neonates who successfully received KMC had a statistically significant increase in weight gain and survival compared to those under standard care [32]. In a survey of those same healthcare workers and caregivers, 88% of respondents were unable to correctly identify eligibility requirements for KMC and over half lacked general knowledge of KMC [33].

Two other studies assessing KMC in the DRC found that KMC was unsupported and rarely practiced at the facility level. An evaluation of 72 rural health facilities found that health facilities were not equipped to support KMC uptake, as only 11% had KMC Kits and an entire province’s health facilities had none [34]. A separate evaluation conducted in 2010 of 53 health facilities in Lubumbashi, DRC found that only 38% (20 facilities) had practiced KMC in the three months preceding the survey [35].

### Caregiver perspective

While research on KMC programs in the DRC has focused on policy, facilities, and health care workers, there is a paucity of research on the caregiver’s perspective. Studies in other low-income countries focused on caregivers and KMC have identified numerous factors associated with uptake [17,36,37]. A systematic review of 112 studies using data from high, middle, and low-income countries on the uptake of KMC found that positive perceptions of the potential benefits of KMC were associated with uptake among caregivers [17]. Societal norms such as beliefs about skin-to-skin contact between infants and their caregivers being inappropriate, and norms against fathers giving direct care to premature infants reduced KMC uptake [14,38]. Communal and familial support were also associated with KMC utilization; there was a decrease in uptake when those close to the mother, specifically her mother-in-law, male partner, and grandmother, did not approve of the practice, and an increase when close female family members supported the mother [39,40]. Peer-to-peer support among mothers on a maternity ward was also positively associated with increased use of KMC [41]. Lack of autonomy in decision-making was a barrier to KMC utilization in Malawi; women mentioned needing their husband’s or mother-in-law’s permission before practicing KMC [42].

To better understand caregiver health intentions and behavior, theories about behavior and action can be used [43,44]. The Integrated Behavioral Model (IBM) was developed from the Theory of Reasoned Action (TRA) and the Theory of Planned Behavior (TPB) and posits that the best predictor of performing a specific behavior is intention (Figure 1) [45,46]. The TRA asserts that the most important determinant of behavior is *behavioral intention* which is influenced by *attitude* and *subjective norms*. The TPB added *perceived control* over behavior as another component in *behavioral intention* [45]. IBM theorizes that intent to perform a specific behavior consists of three components: attitude, perceived norms, and personal agency. Attitude is defined in two ways: (1) experiential attitude, an individual’s emotional response to performing a specific behavior, and (2) instrumental attitude, an individual’s beliefs about anticipated outcomes from performing a specific behavior. Perceived norms consist of both injunctive norms, beliefs about if others approve or disapprove of a behavior and the desire to comply, and descriptive norms, beliefs about if others engage in a specific behavior. Finally, personal agency is defined though perceived control, an individual’s perception of their control over behavioral performance, and self-efficiency, the confidence one has that they can perform a certain behavior to achieve specific outcomes [45]. The IBM has been used to examine intention behind postpartum family planning [47], HIV prevention [45], cervical cancer screening [48], and predict disparities in advance care planning [49]. As the IBM has never been applied to KMC so the impact of intention on practicing KMC is unknown.

**Fig 1.**
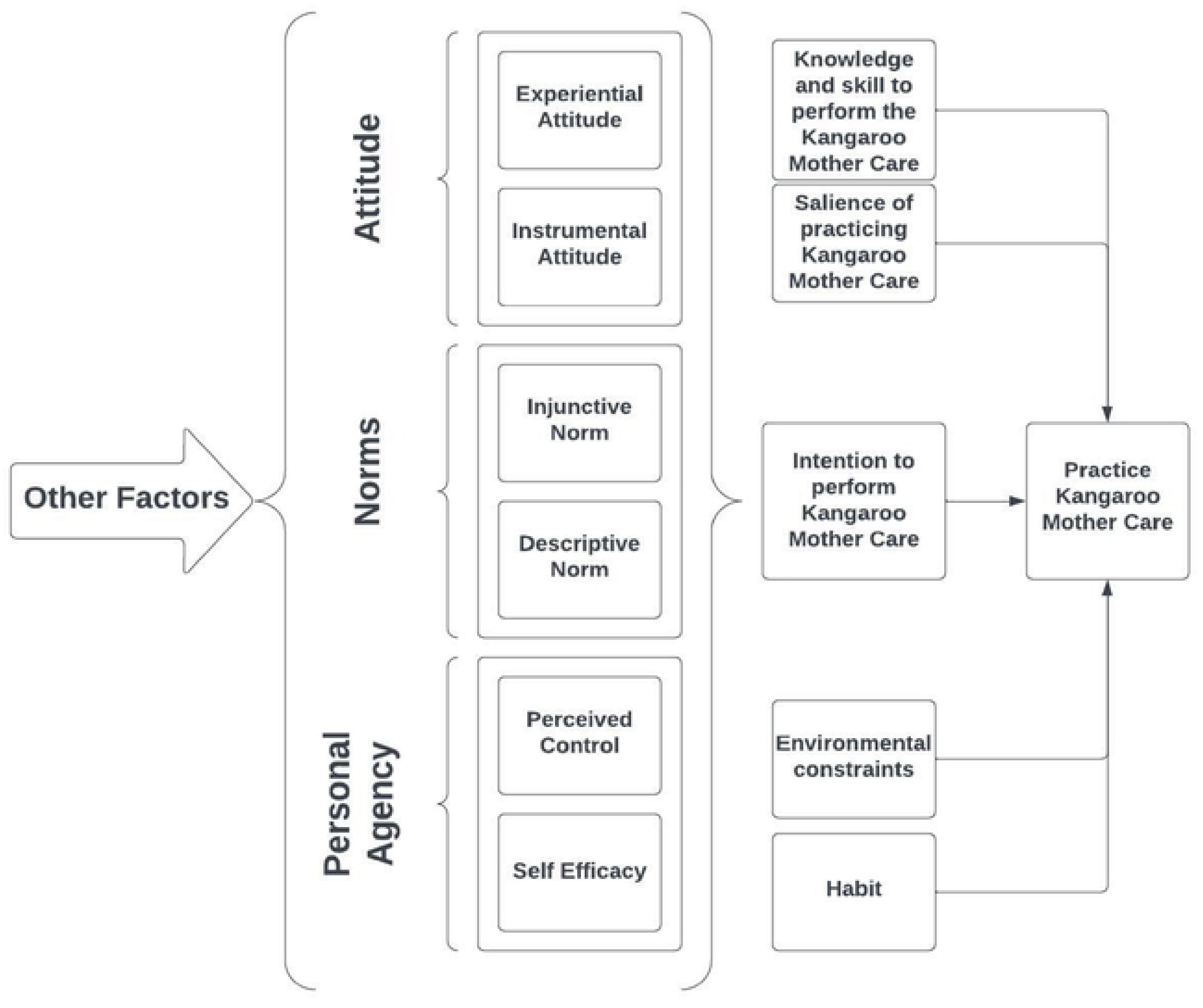
Integrated Behavioral Model of Kangaroo Mother Care. *Experiential attitude:* Belief that KMC intentions or use is associated with specific positive or negative feelings. *Instrumental attitude:* Belief that KMC intentions or use is associated with positive or negative outcomes. *Injunctive norm:* Belief that those most important to the respondent approve or disapprove of KMC and the motivation to comply. *Descriptive norm:* Belief about if other first-time mothers 15-24 years with a LBW or preterm baby practice or intend to practice KMC. *Perceived control:* Belief in one’s control over practicing or intending to practice KMC. *Self-efficacy:* Belief in one’s own the ability to practice KMC.

This paper aims to determine the intention to practice KMC among first-time mothers (FTMs) 15-24 years in Kinshasa, DRC using a modified IBM framework (Figure 2) to explore the associations between KMC intentions and norms, personal agency, and attitudes. A modified IBM framework was used as not all IBM components were measured in the survey. While the IBM model suggests other factors such as knowledge and skill to perform, environmental constraints, and salience of the behavior as influencing behavior along with intention, this paper focuses on the intention component of the model. Secondly, this paper also examines how different demographic characteristics, specifically marital status and age, impact the intention to practice KMC among FTMs 15-24 years in Kinshasa, DRC through an IBM-guided lens.

**Fig 2.**
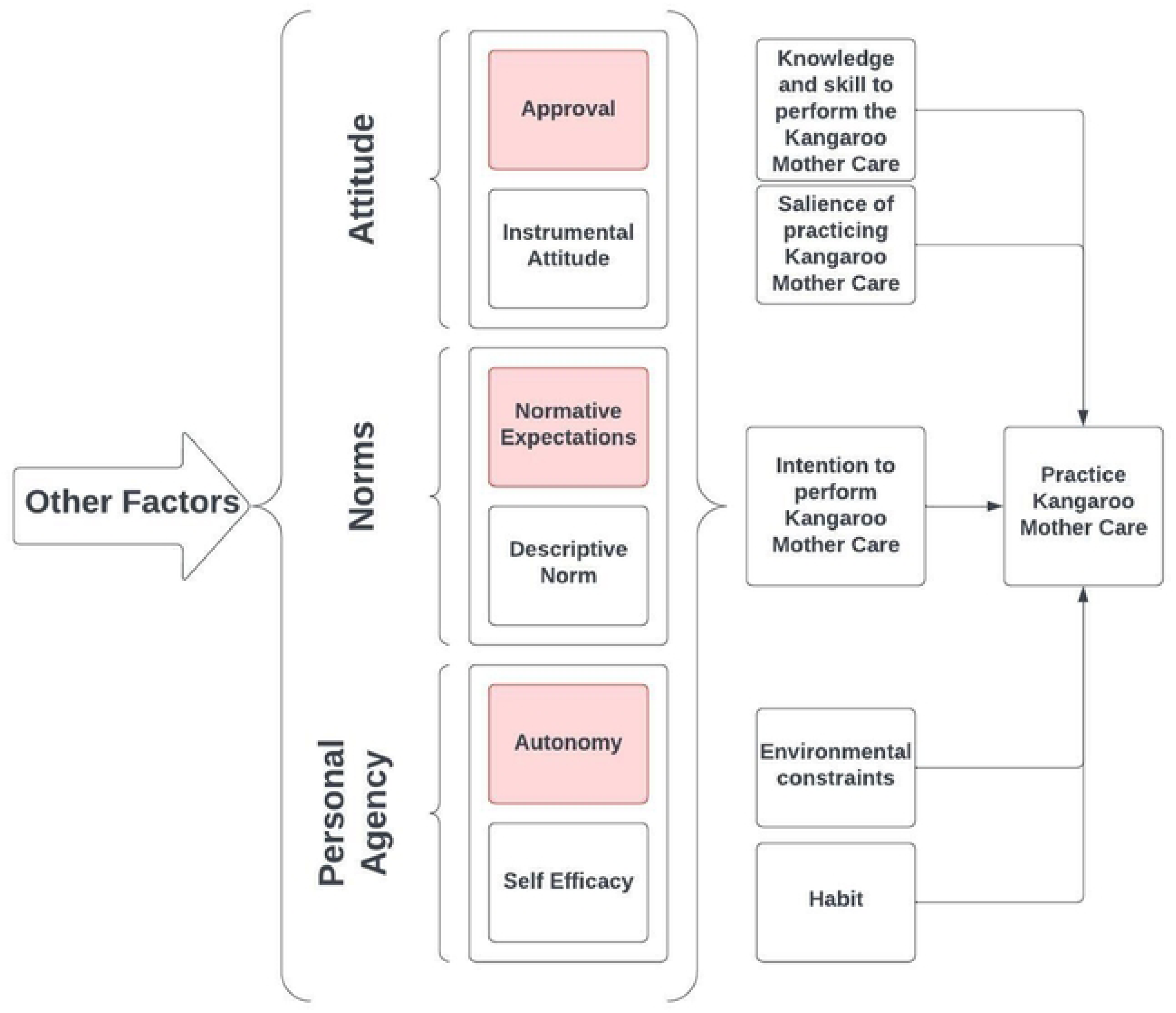
Modified Integrated Behavioral Model of Kangaroo Mother Care. *Approval:* Approval of KMC. *Instrumental attitude:* Belief that KMC intentions or use is associated with positive or negative outcomes. *Normative expectations:* Belief that those most important to the respondent think she ought to practice KMC. *Descriptive norm:* Belief about if other first-time mothers 15-24 years with a LBW or preterm baby practice or intend to practice KMC. *Autonomy:* Belief that the respondent would still practice KMC even if those most important to her did not want her to do it. *Self-efficacy:* General belief in one’s own ability to perform actions to achieve certain outcomes.

This paper tests five hypotheses. The first hypothesis is that descriptive norms and normative expectations are positively associated with the intent to practice KMC. The second hypothesis is that self-efficacy and autonomy are positively associated with the intent to use KMC. As husbands and/or mothers-in-law can be a barrier to KMC uptake by decreasing the mother’s autonomy [42], hypothesis three is that marital status will moderate the association between KMC autonomy and the intent to practice KMC. Since younger age is associated with lower autonomy [50–53], hypothesis four is that age will moderate the relationship between autonomy and KMC intentions. As marital status can influence women’s health behavioral intentions differently depending on their age [54], so the fifth hypothesis is that marital status will moderate the association between age and the intention to practice KMC.

## Methods

### Data

Data were from the 2020 endline survey for Momentum, a Bill & Melinda Gates Foundation funded gender-transformative integrated family planning, maternal and newborn health, and nutrition project that was delivered through nursing students to FTMs 15-24 years and their male partners in Kinshasa, DRC. Data were collected in three intervention health zones (Kingasani, Lemba, and Matete), and three comparison health zones (Bumbu, Ndjili, and Masina1). Nursing students conducted home visits and support group education session with FTMs and their male partners and the home visits included gender-integrated counseling on best practices for newborn health such as KMC. FTMs were recruited at six months pregnant through community outreach and at health facilities. Recruitment took place from September 5 to November 23, 2018 until the desired sample size was reached.

Sample size was estimated based on the likelihood of resorting to cluster sampling if a cohort follow-up study was not possible and was based on detecting a 10-percentage point difference in key behavioral indicators with 99% confidence and 99% power, assuming an attrition rate of 25%, and a design effect of 2.0. A sample size of 1,213 FTMs 15-24 years was estimated for both the intervention and comparison health zones for a total of 2,426 FTMs. The comparison health zones were selected based on similar demographic characteristics to the intervention health zone. FTMs in all health zones were recruited at six months gestation either at health facilities or through community outreach [55]. The survey was administered in 2020 in both intervention and comparison health zones and a total of 1,927 FTMs completed the survey.

The study received approval from the Tulane University Institutional Review Board (1112188) and the Kinshasa School of Public Health Ethics Committee (ESP/CE/066/2018, ESP/CE/08/2020). Written and electronic consent was obtained from all participants before the interview commenced. Both IRB committees waived consent from a parent or legal guardian for FTMs younger than 18 years as some were already married, did not live at home, and were considered to be adults.

### Variables

#### Intention to use KMC

Respondents were asked in the endline survey if they were “very likely,” “likely,” “unlikely,” or “very unlikely” to intend to practice KMC if, in the future, they had a LBW or preterm baby. For this study KMC intentions was measured as a binary variable, with “very likely” defined as intending to practice KMC and coded as “1,” and all other responses coded as “0.” Within the sample 50.7% of respondents were “very likely” to practice KMC.

#### Attitudes

Instrumental attitudes (beliefs that KMC is associated with certain outcomes) were measured by how many benefits of the practice a FTM could name. In the survey KMC was first explained to the FTMs as “a method of care practiced on babies, usually on a low-birthweight or preterm infant, where the infant is held skin-to-skin with his mother, father, or substitute caregiver.” FTMs were then asked, “What do you see as the benefits of Kangaroo Mother Care?” Respondents named up to eight benefits but those who named five or higher were grouped together due to low cell counts and defined as “5 benefits or more.”

#### Approval

KMC approval was measured by asking FTMs if they approved of KMC and responses were recorded as “yes” (1) or “no” (0).

#### Descriptive norms

Descriptive norms were measured using FTMs’ responses to the question on how many other FTMs 15-24 years with a preterm or LBW baby in their community practiced KMC on a five-point scale: all of them, more than half of them, about half of them, less than half of them, or none of them. Responses of “all of them,” “more than half of them,” and “about half of them” were combined and coded as “1” while the remaining categories were coded as “0.”

#### Normative Expectations

Normative expectations are beliefs about what people important to an individual think they ought to be doing. FTMs were asked “Most people who are important to me think I ought to practice Kangaroo Mother Care if I have a low birthweight or preterm baby” and responded on a four-point scale (strongly agree (ref), agree, disagree, strongly disagree). Responses were reverse coded with the highest value representing strong agreement and the lowest representing strong disagreement. Responses for “strongly disagree” and “disagree” were combined due to the low number of “strongly disagree” responses as “disagree/strong disagree.”

#### Self-efficacy

The survey did not ask respondents questions about self-efficacy specific to practicing KMC; however, FTMs were asked questions about general self-efficacy from the General Self-Efficacy Scale [56]. FTMs were asked about ten different self-efficacy behaviors related to overcoming adversity and problem solving, and if the statement was “not true at all,” “hardly true,” “moderately true,” or “always true.” Responses were used to create an additive index for an overall measure of self-efficacy (Cronbach’s alpha of 0.89, range 10-40), with higher values representing higher levels of self-efficacy.

#### Autonomy

Autonomy is the freedom from the external control of others. In this study autonomy was specific to KMC, and FTMs were asked if they would still practice KMC even if those most important to her did not want her to; responses were coded as “yes” (1) or “no” (0).

Additional covariates were marital status, age, education, household wealth, residence in Momentum intervention or comparison health zone, employment status, and previous knowledge of KMC. Information about marital status, age, education, household wealth, residence in an intervention or comparison health zone, and previous knowledge were all from the 2018 Momentum baseline survey as exposure to the project interventions was controlled for in the analysis. Marital status was measured as a binary variable, respondents were coded as “1” if they had never been married “0” (reference category) for all other responses. Age was divided into two groups, 15-19 years (reference group) and 20-24 years. Education was also binary: none/primary/secondary incomplete (reference group) and secondary education complete and higher. Employment status was ascertained by asking FTMs if they had done any work, aside from housework, in the 12 months prior to the baseline survey and responses were recorded as “yes” (1) or “no” (0).

Household wealth was measured as an index created using principal component analysis of variables for the floor, wall, and roofing materials of the dwelling, drinking water, toilet facilities, and ownership of items (radio, television, non-mobile phone, computer, refrigerator, stove, watch, mobile phone, bicycle, motorcycle/scooter, animal-drawn cart, car/truck, a boat with a motor). The index consisted of 19 indicators with a Cronbach’s α of 0.64. The index was divided into terciles of low, medium, and high for ease of interpretation.

Previous knowledge of KMC was measured by asking FTMs if they had ever heard of KMC, and answers were recorded as “yes” (1) or “no” (0). After assessing previous knowledge of KMC, survey enumerators explained KMC to all respondents and all other questions about KMC were asked following this explanation.

### Data Analysis

Descriptive statistics are presented as counts and percentages. The bivariate analysis used Pearson’s chi-square to test the association between the percentage of FTMs who “very likely” intended to practice KMC and attitudes, norms, personal agency, and demographic characteristics. Intent to use KMC was defined using only the “very likely” response because stronger intentions are better predictors of behavior [57]. The data were not weighted as the sample was purposive.

This paper estimated two multivariable logistic regression models. The first model included all variables of interest, and the second model included three interaction terms: the interaction between age and autonomy, the interaction between marital status and autonomy, and the interaction between marital status and age. Multicollinearity was assessed using variance inflation factors (VIF); the highest VIF value was 1.31 and the average was 1.18. Although 1,927 FTMs completed the endline survey, FTMs who were missing information about key demographic characteristics were dropped from the analysis, leaving a study sample of 1,918 FTMs. Statistical analysis was performed using Stata software version 16 (StataCorp, College Station, TX) [58].

## Results

### Participants’ Characteristics

Table 1 shows the demographic characteristics of both the study sample included for analysis and those lost-to-follow up or dropped from analysis. A total of 504 FTMs were lost-to-follow up and nine FTMs were dropped from the analysis due to missing data. Pearson’s chi-square tests found that none of the demographic characteristics differed significantly between those included and those excluded from the study sample except for socioeconomic status.

**Table 1.**
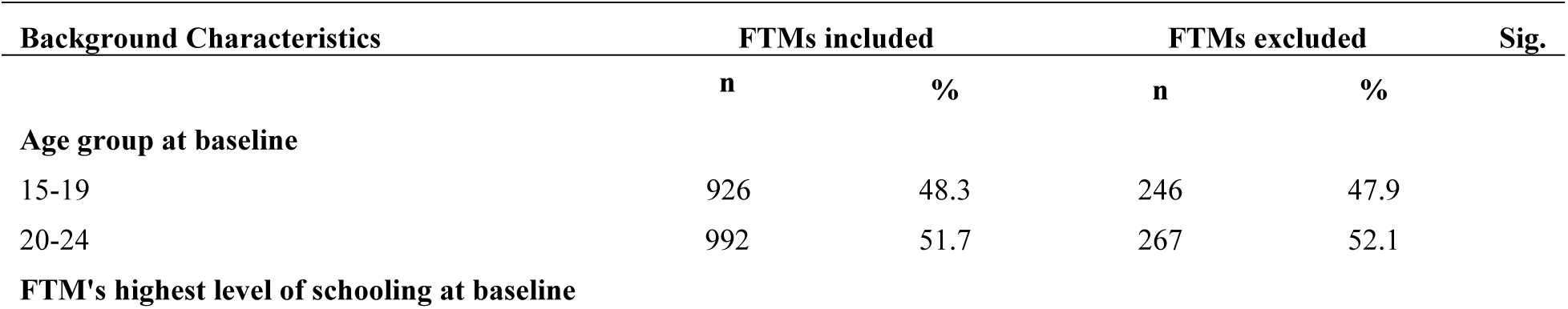
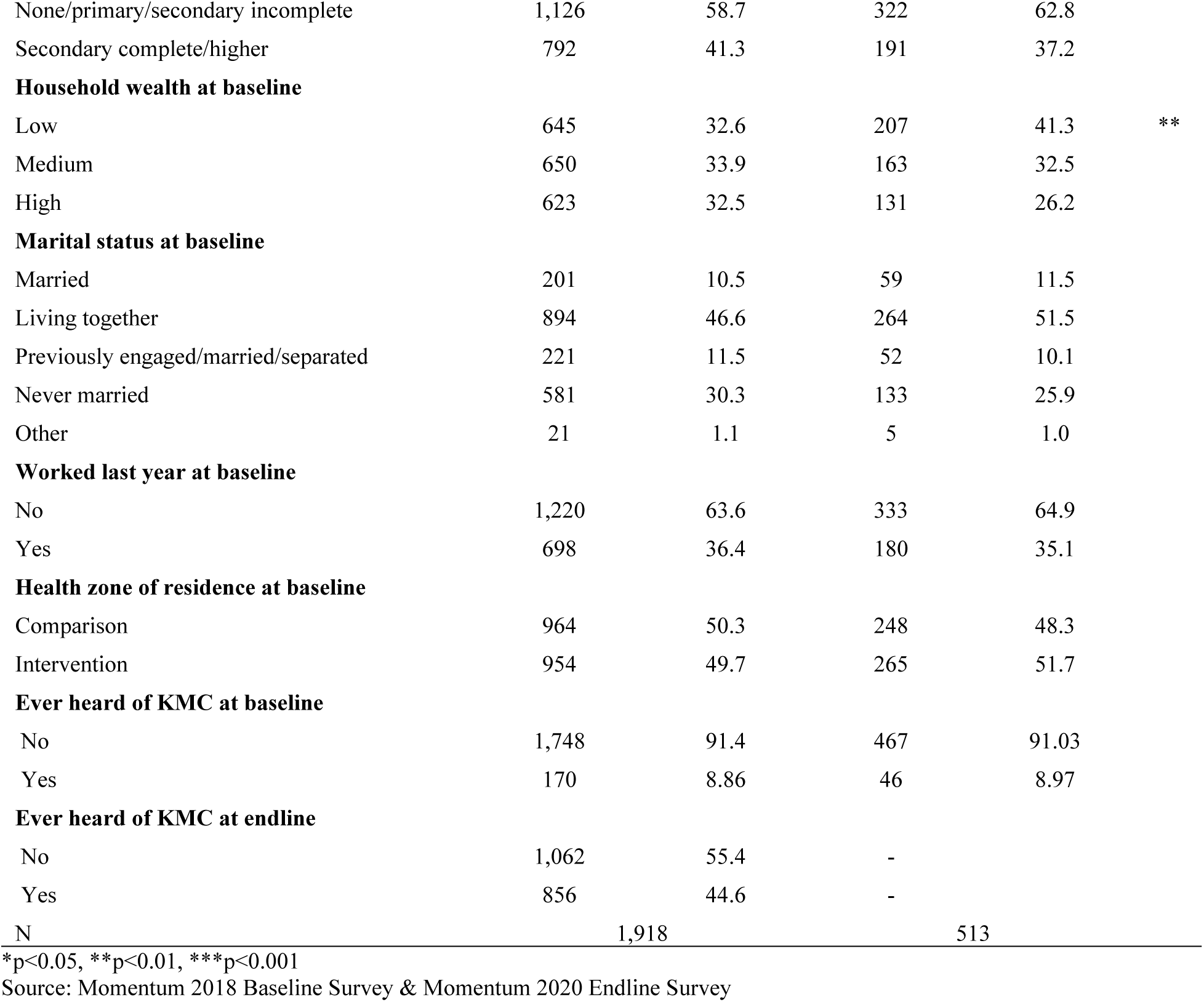
Percent distribution of first-time mothers 15-24 years by inclusion status, according to background characteristics, Kinshasa DRC 2018.

The percentage of FTMs who resided in the poorest households was lower in the included sample than in the excluded sample (33% versus 41%). The mean age at baseline was 20 years for both the included and excluded groups. Among those included, 30% had never been married and were not living with a partner, while 47% were living with a partner, 11% were married, and 12% had been previously married, engaged or were separated. Almost 60% of included FTMs had no schooling, only completed primary school, or had not completed secondary school compared to 63% of excluded FTMs with similar education. About two-thirds of included and excluded FTMs were unemployed the year before baseline. Over 90% of respondents in both the inclusion and exclusion groups had not heard of KMC before at baseline. In the endline survey a little less than half of respondents had heard of KMC (47%).

Table 2 presents the percentage distribution and mean values of the IBM component variables from the endline survey. For attitudes, FTMs could name on average two benefits of KMC (range 0-9) and over 90% approved of KMC. Descriptive norms were low, less than ten percent of FTMs believed that KMC was practiced widely in their community. However, normative expectations were high, with a little less than half of respondents strongly agreeing that those important to them think they should practice KMC. The self-efficacy index mean was 31 with a range from 10 to 40 and almost three-fourths of FTMs were autonomous vis-à-vis KMC.

**Table 2.** Percent distributions and mean index values of explanatory variables, and intentions about KMC of first-time mothers 15-24 years, Kinshasa DRC 2020.

| Explanatory Variables | Total |  |
| --- | --- | --- |
|  | Mean/Percent | Range |
| **Attitude** |  |  |
| Instrumental Attitude | 2.2 (1.2) | 0-9 |
| KMC Approval | 91.7 |  |
| **Norms** |  |  |
| Descriptive norms | 7.7 |  |
| Normative expectations |  |  |
| Strongly disagree/Disagree | 13.4 |  |
| Agree | 40.8 |  |
| Strongly agree | 45.8 |  |
| **Personal agency** |  |  |
| Mean self-efficacy index | 30.8 (6.1) | 10-40 |
| KMC autonomy | 71.3 |  |
| **KMC intentions** |  |  |
| Strongly intend to practice KMC | 50.7 |  |
| N | 1,918 |  |

() Standard deviation
Source: Momentum 2020 Endline Survey

### Bivariate Tabulations

Table 3 shows the percentage of FTMs who intend (“very likely”) to practice KMC if they had a preterm or LBW baby by explanatory variables. Overall, half of FTMs stated that they intended to practice KMC (51%). All the IBM components were significantly associated with the percentage of FTMs who intend to practice KMC. For FTMs who knew no KM benefits, only one-third were very likely to practice KMC, while about half of FTMs who could name one to four benefits intended to practice KMC. FTMs who could name five or more benefits of KMC had the highest proportion of “very likely” KMC users. Among FTMs who approved of KMC, a little more than half (56%) were very likely to practice KMC, while only 4% of those who disapproved were.

**Table 3.** Percentage of first-time mothers 15-24 years who were very likely to intend to practice KMC and control variables.

| Variables | Very likely to practice KMC |  |
| --- | --- | --- |
|  | % | N |
| **Attitude** |  |  |
| Instrumental Attitude | *** |  |
| No KMC benefits | 36.7 | 90 |
| 1 KMC benefit | 42.7 | 328 |
| 2 KMC benefits | 54.2 | 868 |
| 3 KMC benefits | 53.3 | 409 |
| 4 KMC benefits | 47.1 | 157 |
| 5+ KMC benefits | 57.6 | 66 |
| KMC Approval | *** |  |
| Yes | 55.5 | 1,758 |
| No | 4.4 | 160 |
| **Norms** |  |  |
| Descriptive norms | *** |  |
| Half to all | 81.1 | 148 |
| Less than half | 48.2 | 1,770 |
| Normative expectations | *** |  |
| Strongly agree | 85.5 | 878 |
| Agree | 24.0 | 782 |
| Strongly disagree/Disagree | 13.2 | 258 |
| **Personal agency** |  |  |
| Self-efficacy | *** |  |
| Low | 37.4 | 672 |
| Medium | 58.7 | 637 |
| High | 57.1 | 609 |
| KMC autonomy | *** |  |
| Yes | 54.1 | 1,367 |
| No | 42.5 | 551 |
| **Age group** |  |  |
| 15-19 | 49.8 | 926 |
| 20-24 | 51.6 | 992 |
| **Never Married** |  |  |
| No | 50.5 | 1,337 |
| Yes | 51.3 | 581 |
| **FTM's highest level of schooling** |  |  |
| None/primary/secondary incomplete | 49.4 | 1,126 |
| Secondary complete or higher | 52.7 | 792 |
| **Household wealth** |  |  |
| Low | 49.5 | 645 |
| Medium | 50.3 | 650 |
| High | 52.5 | 623 |
| **Worked last year** |  |  |
| No | 52.3 | 1,220 |
| Yes | 48.0 | 698 |
| **Previous knowledge of KMC** |  |  |
| No | 50.5 | 170 |
| Yes | 53.5 | 1,748 |
| **Intervention health zone** |  |  |
| Comparison | 49.4 | 964 |
| Intervention | 52.1 | 954 |
| Total | 50.7 | 1,918 |

\*p<0.05, \*\*p<0.01, \*\*\*p<0.001
Source: Momentum 2018 Baseline Survey and 2020 Endline Survey

For descriptive norms, the proportion of FTMs who thought more than half or half to all FTMs in their community with a LBW or preterm baby practiced KMC was one and half times larger than those who believed less than half of FTMs in their community practiced KMC. Three times as many FTMs responded that they intended to practice KMC if they strongly agreed that most people important to them thought they ought to practice the method compared to those who just agreed. The proportion of FTMs who strongly agreed was six times the proportion of FTMs who disagreed. The lowest self-efficacy scale category had the lowest prevalence of FTMs who intended to practice KMC (37%), while the medium and high categories were similar (59% vs. 57%). FTMs who were autonomous had a higher prevalence of FTMs who intended to practice KMC (50%) compared to those who were not (43%). None of the demographic variables were significantly associated with being “very likely” to intend to practice KMC.

### Multivariable Analysis

Table 4 presents the results for the multivariable logistic regression analysis on the association between the intent to practice KMC and our adapted IBM constructs, after controlling for baseline socioeconomic characteristics. Model 1 was the base model and Model 2 included interaction terms for KMC autonomy and marital status, KMC autonomy and age, and age and marital status.

**Table 4.**
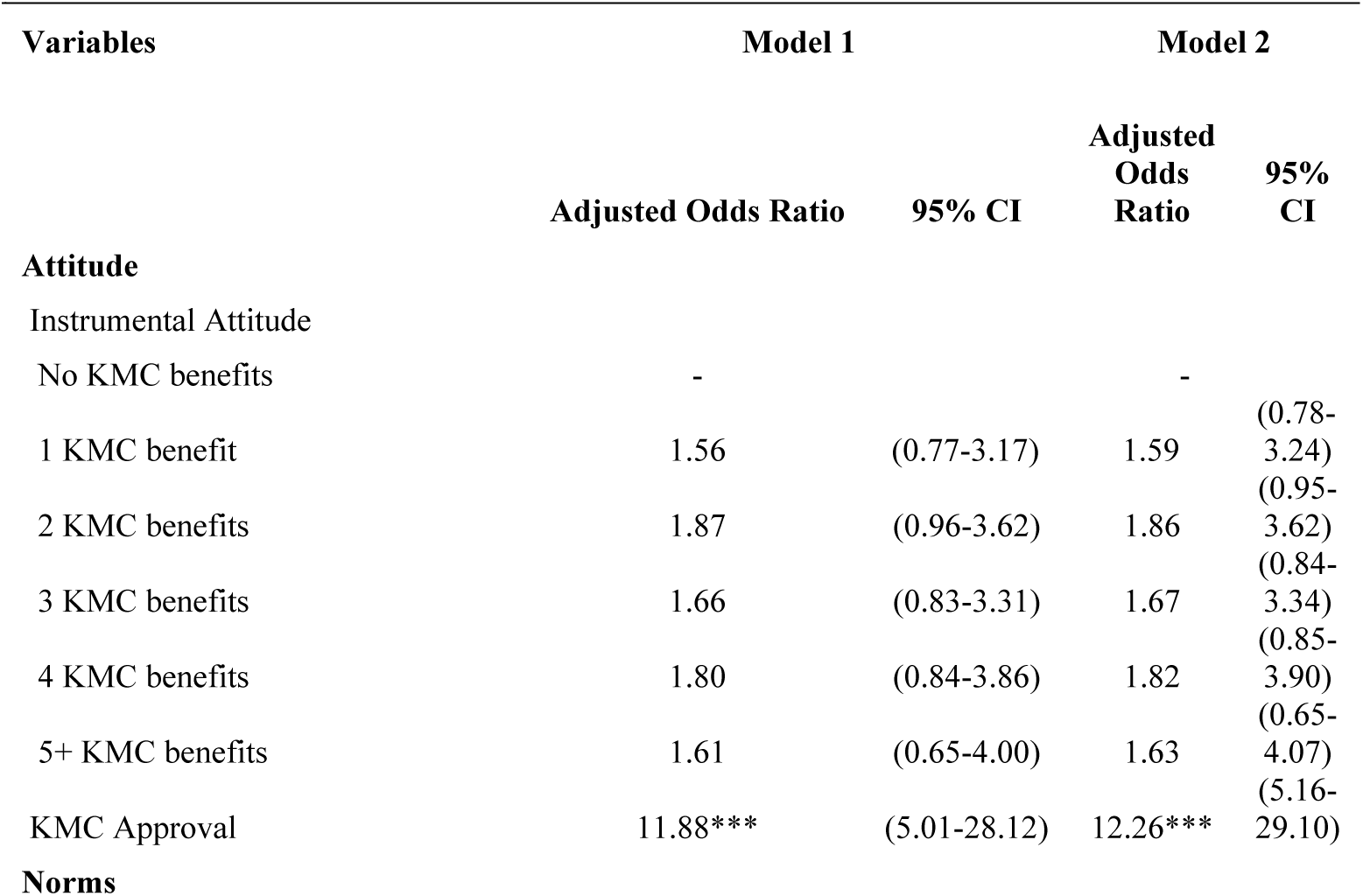
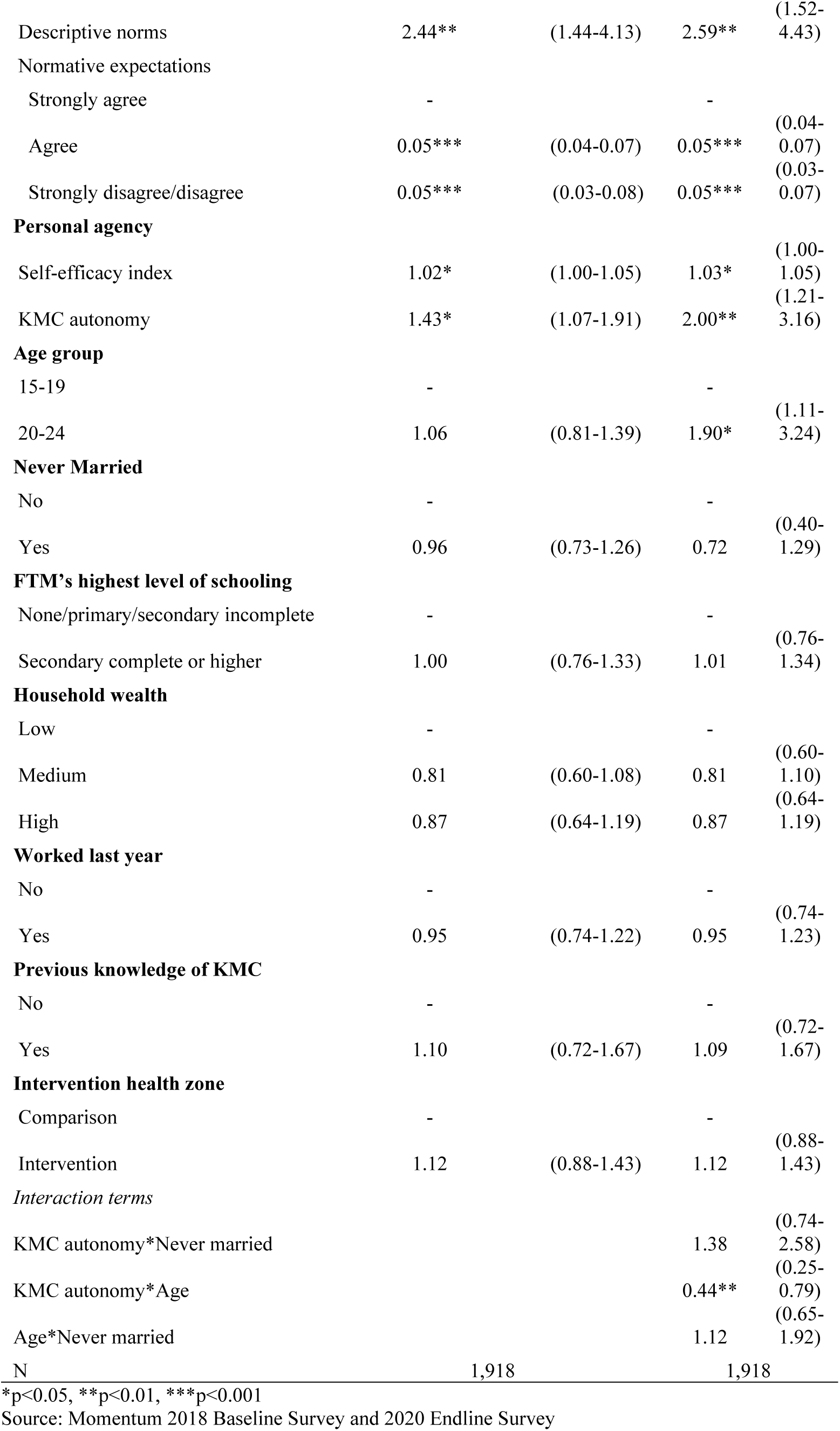
Multivariable logistic regression models of Kangaroo Mother Care intentions among first-time mothers 15-24 years, Kinshasa, DRC 2020.

In Model 1, all the IBM components were significantly associated with the intent to practice KMC except for instrumental attitudes. Those who approved of KMC had almost 12 times higher odds (95% CI= (5.01-28.12)) of intending to practice KMC compared to those who did not. Descriptive norms (perceptions about KMC utilization among FTMs 15-24 years in the community) were significantly and positively associated with the intent to practice KMC. This finding suggests that FTMs who believed that at least half of FTMs 15-24 year in their community practiced KMC were more likely to intend to use the practice themselves (OR=2.44; 95% CI= (1.44-4.13)). For normative expectations, compared to those who strongly agreed, those who disagreed/strongly disagreed (OR=0.05; 95% CI= (0.03-0.08)) and those who agreed (OR=0.05; 95% CI=(0.04-0.07)) were statistically significantly less likely to intend to practice KMC.

For the personal agency component of IBM, both self-efficacy and KMC autonomy were statistically significant, meaning that those who rated themselves highly on the general self-efficacy scale and those who would still practice KMC even if those important to them did not want them to, were more likely to intend to practice KMC. The odds of intending to perform KMC increased by 2% (95% CI=(1.00-1.05)) for each increase in the self-efficacy index while those who were autonomous vis-à-vis KMC had about 40% increased odds (95% CI=(1.07-1.91)) of stating they were very likely to practice KMC.

None of the coefficients for the number of KMC benefits known were significant compared to knowing no benefits, suggesting that instrumental attitudes do not influence the intent to practice KMC. A sensitivity analysis testing the association between intent to practice KMC and naming any benefit of KMC also found no significance, as did knowledge of two (the mean) or more benefits.

The coefficient for KMC approval had a large confidence interval, indicating that the estimate was imprecise; this was likely due to small cell counts as approval of KMC was over 90%. Due to lack of variability in the KMC approval variable, the regression was run excluding it from the analysis, but the coefficients for instrumental norms, normative expectations, and autonomy changed by more than 10%, indicating that KMC approval was a confounding variable and should be kept in the model.

The findings from Model 2 were the same as Model 1 except the estimate for age was significant. FTMs 20-24 years had 90% increased odds (95% CI=(1.11-3.24)) of practicing KMC compared to FTMs 15-19 years. Of the interaction terms, only the interaction between KMC-specific autonomy and age was significant (OR=0.44; 95% CI=(0.25-0.79)), indicating that age moderates the relationship between autonomy and intention to practice KMC. The marginal effects of the interaction showed that while the probability of KMC intentions for older FTMs 20-24 years were similar between autonomy levels, the probability of intending to practice KMC increased significantly between the low and high autonomy for FTMs 15-19 years. The probability of intending to practice KMC was slightly higher for younger autonomous FTMs compared to older autonomous FTMs. Both the interaction between marital status and KMC autonomy and the interaction between age and marital status were not significant, indicating that the association between the intent to practice KMC and KMC autonomy did not differ by marital status and marital status did not moderate the association between age and KMC intentions. Besides age, none of the other demographic characteristics were significantly associated with the intention to practice KMC in either of the models.

To better examine differences by age, Model 2 was disaggregated by age group (see Supplemental Table 1) and the interaction between KMC autonomy and marital status was significant for FTMs 20-24 years. The marginal effects of the interaction showed that while FTMs who were never married had little difference in the probability of intending to practice KMC between autonomy levels, FMTs who were currently/previously married had a decrease in probability of the outcome when they were autonomous. For FTMs 15-19 years the confidence intervals for many of the estimates were large, making interpretation unreliable as the estimates were not precise.

A sensitivity analysis was performed redefining the dependent variable to include both “very likely” and “likely” to intend to perform KMC. Using this definition 87.3% of respondents intended to practice KMC (See Supplemental Table 2). Analysis with the newly defined dependent variable found that both descriptive norms and the general self-efficacy index were no longer significant in either model. In Model 4, knowing two benefits of KMC was significant and positively associated with KMC intentions compared to knowing no benefits, though no other number of KMC benefits known was significant. The interaction term between KMC autonomy and marital status was significant in Model 5, indicating that marital status moderated the association between autonomy and the intent to practice KMC. Analyzing the marginal effects of the interaction showed that while there was no statistical difference in the probability of intending to practice KMC among high autonomy FTMs, for low autonomy FTMs those who were never married had a statistically higher probability of intending to practice KMC compared to FTMs who were currently or previously married. Many of the odds ratio estimates had wide confidence intervals, most likely due to a lack of variability in both the dependent and independent variables.

As the neonates of adolescent mothers (15-19 years) have higher rates of adverse health outcomes, models were analyzed disaggregating by age[59]. Disaggregating by age showed that the significance of the modified IBM components differed by age group (Supplemental Table 1). While normative expectations and KMC approval were statistically significant for both age groups, neither the self-efficacy index nor descriptive norms were statistically significant for FTMs 20-24 years. The interaction term between KMC autonomy and marital status was significant for FTMs 20-24 years. While autonomous older FTMs who had never been married had a higher probability of intending to practice KMC compared to non-autonomous older FTMs, the opposite relationship was found among currently/previously married FTMs 20-24 years.

## Discussion

This is the first study to examine KMC intentions among adolescent and young first-time mothers using the IBM framework in a low-resource setting. Overall, the models showed support for the modified IBM, finding that norms (descriptive norms and normative expectations), personal agency (self-efficacy and autonomy), and attitudes (approval) were positively associated with KMC intentions.

The only IBM variable that was not significantly associated with KMC intentions was instrumental attitudes, which measured the respondent’s perceived benefit of KMC practice. This finding was in contrast to previous research which found that positive perceptions of potential benefits of KMC among mothers, fathers, and their families supported uptake [17,39]. This finding could be due to differences in study populations; most studies on KMC uptake were not focused on young first-time mothers and instrumental attitudes could be less impactful in this group. Previous research on instrumental attitudes among adolescents in Europe found that text messages using instrumental messages were not as effective in promoting healthy eating or physical activity as affective messages [60,61]. Additionally, these studies used uptake as the outcome variable, not intent, so, as a matter of conjecture, it may be that instrumental attitudes are associated with uptake of the method but not the intention to practice.

Findings from Model 2 showed that age was positively associated with the intent to practice KMC; FTMs 20-24 years had nearly double the odds of KMC intentions compared to FTMs 15-19 years with the same level of autonomy. The significant interaction between autonomy and age showed that the association between autonomy and the intent to practice KMC was moderated by age. Analysis of the marginal effects found that the probability of intending to practice KMC did not change by autonomy status for FTMs 20-24 years, but FTMs 15-19 years had a significant increase in the probability of intending to practice KMC from the low to high level of autonomy. These results indicate that increasing autonomy among younger FTMs could have a greater impact on deciding to practice KMC compared to older FTMs.

When disaggregating models by age, the probability of KMC intentions was lower among autonomous currently/previously married FTMs compared to non-autonomous ones. This finding could indicate that strategies for increasing KMC intentions among older FTMs may need to differ by marital status and that effort to increase autonomy in this population may only benefit never married FTMs.

This paper’s findings support four of the five hypotheses and contribute to the literature in several ways. First, this paper examined the association between norms, personal agency, and attitudes and KMC quantitatively while the majority of studies examining these drivers and caregiver uptake are qualitative [26,39,42,62]. Second, this study focused on a vulnerable population, first-time mothers 15-24 years, whose neonates are at higher risk of adverse outcomes and therefore could benefit greatly from the practice. Third, this paper highlighted several possible methods to drive community demand for KMC among this vulnerable population. Finally, this study adds to the limited research on KMC in the DRC. With high rates of neonatal mortality and morbidity, and 43.2% of neonatal deaths due to preterm child deaths, the importance of understanding the applicability of a low-cost but effective method such as KMC is a key step in reducing these adverse outcomes[63].

Findings from this paper complement the health facility-based work being carried out by the USAID-funded Integrated Health Project (IHP), with implementing partner Abt Global [64], in health facilities in the southwest of the DRC. While the IHP focuses on supply-side barriers, this study provides evidence for the creation of demand centered programs on KMC among FTMs 15-24 years. As over 90% of respondents delivered at a health facility, a facility-based KMC strategy fits this population. KMC units within health facilities in which mothers can initially practice KMC alongside their peers could demonstrate to the mother that KMC is practiced within the community (descriptive norms) as well as provide privacy. KMC units also can increase socialization for the mother while she practices the method, as a common critique of KMC was that the method could be both isolating and boring [41,62].

The significance of normative expectations and autonomy suggest that community engagement in KMC must extend beyond the mother to her partner, extended family, and the wider community. While KMC is a low resource intervention, performing KMC can be labor intensive and cumbersome for the caregiver [17]. Women in the DRC are primarily responsible for household duties [65] and family pressure to continue to perform these tasks after birth can be a barrier to practicing KMC [40]. In other low-income settings KMC uptake was increased when female relatives of the mother were able to help with household work, allowing the mother time to practice KMC [14]. Additionally, having multiple caregivers practice KMC prevents caregiver burnout and alleviates fatigue [66]. Interventions should address determinants simultaneously as multiple components were significant.

### Limitations

This study has some limitations that should be considered when evaluating the results. First, this study used a cross-sectional non-experimental design. Due to non-randomized selection, there may be some uncontrolled confounders and results are not generalizable. Due to the cross-sectional design, only correlations could be determined.

Another limitation was that 91% of FTMs at baseline and 55% of FTMs at endline reported that they had not heard of KMC. However, the IBM framework posits that knowledge is not necessary for determining intention, as it only influences utilization. While an explanation of KMC was given to FTMs by enumerators during the survey, there likely was some social-desirability bias in FTMs’ responses. A sub-analysis of only those who reported that they had heard of KMC in the endline survey found the same results as the full analysis except that self-efficacy and KMC autonomy were no longer significantly associated with the outcome (Supplemental Table 2). Additional analysis using multivariate logistic regression in which knowledge of KMC was the dependent variable and intent to practice KMC was an independent variable found that while knowledge was associated with all the IBM components, it was not associated with the intent to practice KMC. Review of the literature found that while knowledge of a behavior and the intent to perform the behavior were associated, they were mediated by variables such as norms, attitudes, and personal agency [67–69]. While measuring mediation by IBM components on the relationship between knowledge and intent is outside of the scope of this current study, future research examining this relationship may be helpful. One paper based in Ethiopia found an association between knowledge of the COVID-19 vaccine and the intention to be vaccinated [70], but there is no evidence that this relationship can be applied to KMC. Finally, in Iran, a KMC educational intervention based on improving the components of the Theory of Planned Behavior (attitude, subjective norms, and perceived behavioral norms), found an increase in the intention to practice KMC [71]. While the study found a significant difference in mean knowledge of KMC in the intervention group before and after the intervention, the authors did not test the relationship between overall knowledge of KMC and the intention to perform KMC [71].

For both the questions about normative expectations and autonomy, there was no information about who was most important to the FTM. Those who were most important to the FTM could differ by question and the survey did not capture that difference. Peer groups could exert more influence on younger populations than family, and those who influence the FTM could differ by the FTM’s marital status. Additionally, for descriptive norms, FTMs estimated the use of KMC among their peers which may not reflect actual use. Social norms can be misperceived, and this misperception could impact KMC intentions. Some variables, such as KMC approval and descriptive norms, lacked variability and had large confidence intervals. These variables were included, however, to control for confounding or for fidelity to the theoretical framework.

This paper used a modified IBM framework; experiential attitudes (attitudes about how an action or behavior will feel), injunctive norms (perceived referent approval or disapproval of KMC), and perceived control (how much control someone perceives they have over a certain behavior) were not included. Instead of experiential attitudes about KMC, approval for KMC was used as a second measurement of attitude. While both indicators examine positive feelings towards KMC, they measure two distinct concepts. Normative expectations, what those most important to the FTM think she should do, were included instead of injunctive norms. Although these concepts are related, normative expectations examine what others think an individual ought to be doing while injunctive norms explore the approval or disapproval of a specific behavior by others.

This study included women’s autonomy in the model instead of perceived control. The autonomy variable measured FTMs’ desire to perform KMC despite disapproval from others, while perceived control is defined as FTMs’ perception of the ease or difficulty of performing a specific behavior [72]. Though autonomy could be considered a component of perceived control, they are not synonymous concepts; perceived control accounts for the ability to overcome barriers other than the influence of others. The self-efficacy measure was not specific to KMC but a general measure of self-efficacy. While a more specific measurement of self-efficacy to KMC may predict intentions more precisely, it is not clear that general perceptions of self-efficacy would change substantially when applied to performing a specific activity such as KMC.

Additionally, the measure for instrumental attitudes did not capture the entirety of the concept. Instrumental attitudes were only measured by respondents naming potential benefits of practicing KMC and anticipated negative consequences were not included as they were not collected in the survey. While only positive consequences were measured, the findings still provide useful information in that positive perceptions of practicing KMC were not sufficient for impacting KMC intentions among the sample population.

Finally, intent to practice KMC does not necessarily translate into actual practice. The IBM posits that uptake depends on several factors in addition to intention such as the knowledge and skill to perform the behavior, salience of the behavior, environmental constraints, and habit. The scope of this paper was limited to the intention component of the IBM since, as the model suggests, intentions are a good indicator of acceptability and a necessary predictor of uptake. However, interventions to increase uptake must not only consider factors behind intention, but also other relevant determinants. For example, as per the IBM, knowledge and skill are prerequisites for uptake, not intention, and unprompted knowledge of KMC (measured by whether the participant had ever heard of KMC) was low in the sample. Unless women in the DRC are made aware of the method and are taught the skills to practice it, uptake of KMC will likely be low. Actual practice of KMC could not be included in this study as the number of FTMs who reported having a LBW was small, less than 10% of the sample, and only those who reported having a LBW were asked if they practiced KMC.

## Conclusion

The effectiveness of KMC in low resources settings in reducing neonatal morbidity and mortality, improving the infant-caregiver bond, and enhancing the newborn’s immunity has been repeatedly established [14–17], however, implementation in the DRC has floundered [17,37,40]. Global interest in KMC is high, with the WHO releasing an implementation strategy guide for scaling up KMC in different country contexts in early 2023 [73]. The DRC can take advantage of global enthusiasm for KMC and develop programs to support adolescent and young mothers in adopting KMC using community-based approaches similar to the nursing-student model used by the Momentum Project. A community-based strategy based on an IBM framework to increase awareness and acceptance of the practice coupled with the facility-based work already being carried out in the DRC could help positively impact the neonatal mortality and morbidity trends in the country.

## Data Availability

The datasets used and/or analyzed during the current study are available from the corresponding author (in French) on reasonable request.

## List of abbreviations

ANC: antenatal care
CI: confidence interval
DRC: Democratic Republic of Congo
ENAP: Every Newborn Action Plan
FTM: first-time mother
HMIS: health management information systems
IBM: Integrated Behavioral Model
IHP: Integrated Health Project
KMC: kangaroo mother care
LBW: Low birth weight
LMIC: Low- and middle-income country
TPB: Theory of Planned Behavior
TRA: Theory of Reasoned Action
USAID: United States Agency for International Development
WHO: World Health Organization

## Acknowledgements

The authors would like to acknowledge the Bill and Melinda Gates Foundation (grant number INV_009536), the Momentum study participants, the study enumerators and all DRC partners who worked on this project.

---

**Supplemental Table 1. Logistic regression models of KMC intentions by age group, FTMs 15-24 years, Kinshasa, DRC 2020**

**Supplemental Table 2. Logistic regression models of KMC intentions (“very likely,” “likely”), FTMs 15-24 years, Kinshasa, DRC, 2020**

